# White matter integrity is associated with cognition and amyloid burden in older adult Koreans along the Alzheimer’s disease continuum

**DOI:** 10.1101/2023.04.05.23288147

**Authors:** Lauren Rose Hirschfeld, Rachael Deardorff, Evgeny J. Chumin, Yu-Chien Wu, Brenna C. McDonald, Sha Cao, Shannon L. Risacher, Dahyun Yi, Min Soo Byun, Jun-Young Lee, Yu Kyeong Kim, Koung Mi Kang, Chul-Ho Sohn, Kwangsik Nho, Andrew J. Saykin, Dong Young Lee, the KBASE Research Group

## Abstract

**BACKGROUND:** White matter (WM) microstructural changes in the hippocampal cingulum bundle (CBH) in Alzheimer’s disease (AD) have been described in cohorts of largely European ancestry but are lacking in other populations.

**METHODS:** We assessed the relationship between CBH WM integrity and cognition or amyloid burden in 505 Korean older adults aged ≥55 years, including 276 cognitively normal older adults (CN), 142 mild cognitive impairment (MCI), and 87 AD, recruited as part of the Korean Brain Aging Study for the Early Diagnosis and Prediction of Alzheimer’s disease (KBASE) at Seoul National University.

**RESULTS:** Compared to CN, AD and MCI subjects showed decreased WM integrity in the bilateral CBH. Cognition, mood, and higher amyloid burden were also associated with poorer WM integrity in the CBH.

**CONCLUSION:** These findings are consistent with patterns of WM microstructural damage previously reported in non-Hispanic White (NHW) MCI/AD cohorts, reinforcing existing evidence from predominantly NHW cohort studies.

## BACKGROUND

Alzheimer’s disease (AD) is the most common form of dementia, characterized by hallmark neuropathologies including extracellular amyloid plaques and intracellular neurofibrillary tau tangles, which are thought to lead to downstream neurodegeneration [1]. Initial pathological damage is generally seen in the medial temporal lobe (MTL), primarily affecting the entorhinal cortex and hippocampal regions, which are crucial for memory formation and recall [2]. Clinically, AD is characterized along a diagnostic continuum, where preclinical disease processes, including amyloid deposition, may contribute to progression from cognitively normal (CN) status to mild cognitive impairment (MCI), and/or AD dementia [3].

The lack of inclusiveness and diversity in clinical research on AD significantly limits the generalizability of existing findings. This important limitation stems from broader issues of stigma, socioeconomic disadvantages, and health inequities impacting underrepresented populations [4, 5]. For example, the United States Veterans Health Administration reported that the incidence of dementia among older adults was disproportionately high in minority groups (Asian, African American, and Hispanic) compared to non-Hispanic White (NHW) participants [6]. The genetic architecture of AD risk and resilience may also differ more than is currently known across ethnoracial populations, as recent genome-wide association studies indicate the possibility of differential genetic contributions to AD in East Asian compared to European/ NHW ancestry [7]. Thus, it is important to examine large-scale cohorts of races/ethnicities other than those of European ancestry to fully characterize similarities and differences in the development and progression of AD.

A number of studies have focused on the role of white matter (WM) microstructure in early AD processes. WM consists of axons surrounded by myelin, a fatty lipid sheathing that insulates axons for optimal signal conduction. The dysfunction and degeneration of WM has been implicated in AD pathogenesis [8-10]. A major WM tract known as the cingulum connects the frontal, medial temporal, and parietal cortices [11, 12] and is a critical part of the Papez Circuit, a group of limbic regions including the hippocampus that are thought to contribute to memory function [12]. Previously, impaired connectivity in the Papez Circuit was demonstrated in CN *APOE* ε4 allele carriers who are at increased risk of developing AD and related amyloid pathology [13]. Additional studies from the Pre-Symptomatic Evaluation of Experimental or Novel Treatments for AD (PREVENT-AD) and Dominantly Inherited Alzheimer’s Disease Network (DIAN) have implicated the posterior cingulum as impaired in early-onset dominantly inherited AD, where disease is caused by one or more autosomal dominant mutations in the *PSEN1, PSEN2*, or *APP* genes [14]. The cingulum bundle is also directly associated with the default mode network (DMN) [15], and impairments in the DMN have been consistently described in AD [16, 17]. Further, the cingulum and Papez Circuit are thought to play a role in depression [11, 18-20], which has been previously identified as a major risk factor for AD in both White [21, 22] and Korean populations [23].

The cingulum bundle of the hippocampus (CBH), a posterior region of the cingulum [24] with proximity to the hippocampus, has been implicated in memory recall ability in CN adults [25]. Studies using diffusion tensor imaging (DTI), which measures the diffusion of water molecules, can provide information about WM microstructure [26]. Prior DTI studies have established a role for the microstructural integrity of the cingulum, and especially the CBH, in memory and cognitive impairment related to dementia. Previously, lower fractional anisotropy (FA) and higher radial diffusivity (RD) of the parahippocampal cingulum (indicative of reduced WM integrity), have been associated with the presence of an MCI [27-32] or AD diagnosis [31-34] in studies of predominantly NHW populations. Further, higher amyloid burden, measured through Positron Emission Tomography (PET) with the beta-amyloid tracer [^11^C]-labeled Pittsburg Compound-B (PiB), was associated with lower FA in the parahippocampal cingulum longitudinally, though the same study did not find a direct relationship between *APOE* ε4 allele positivity and FA in parahippocampal WM [35].

A smaller number of studies have evaluated these metrics in non-White populations. For example, another diffusion imaging technique, diffusion spectrum imaging (DSI), demonstrated an association between impaired cognition, especially memory, and lower left cingulum bundle FA in sporadic early onset AD in a small Taiwanese cohort (CN n=15, MCI n=8, AD n=9) [36]. Additionally, findings in a small cohort of Korean individuals (CN n=18, MCI n=19, AD n=19) recruited at Seoul National University (SNU) also demonstrated decreased FA of the parahippocampal cingulum in MCI and AD patients compared to controls, which was associated with episodic memory function [29]. Taken together, these studies implicate disrupted integrity of parahippocampal cingulum WM microstructure as a possible early pathological marker of AD.

Though recruitment strategies for enhancing diversity have been implemented [37, 38], there is still a disparity in cohort size seen in studies of predominantly NHW cohorts and other ethnicities, including Asians. To that end, our goal was to investigate WM integrity of the CBH in a large single site cohort of older Koreans along the AD continuum (CN n=276, MCI n=142, AD n=87), as well as the association of this region with cognition and amyloid deposition. First, we assessed diagnostic group differences in four DTI scalar measures from the CBH, including FA, mean diffusivity (MD), axial diffusivity (AxD), and RD. We additionally investigated associations of these WM integrity metrics with multiple tests of memory and mood to further explore the relationship between the parahippocampal cingulum and cognition in patients along the AD continuum. Finally, we examined [^11^C]PiB-PET standardized uptake value ratio (SUVR) in the MTL, where the hippocampus is located, to investigate the relationship between amyloid deposition and WM integrity in this region.

## MATERIALS AND METHODS

### Participants

Participant demographics can be found in **Table 1**. The cohort was chosen from data collected as part of the initial phase of the Korean Brain Aging Study for the Early Diagnosis and Prediction of Alzheimer’s Disease (KBASE) with approval by the Institutional Review Boards of Seoul National University (SNU) Hospital and SNU-SMG Boramae Medical Center and in accordance with the Declaration of Helsinki as previously described [39]. Briefly, participants ages 55-90 from KBASE included (1) CN older adults with a Clinical Dementia Rating (CDR) of 0 (n=276), (2) older adults with MCI with self, informant, or clinician-based memory complaints, objective memory impairment, a global CDR score of 0.5, and performance on at least one of four episodic memory tests at least one standard deviation below age, sex, and education adjusted norms (n=142), and (3) older adults diagnosed with probable AD dementia in accordance with the Diagnostic and Statistical Manual of Mental Disorders 4^th^ Edition (DSM-IV-TR) and NIA-AAA guidelines with a global CDR Score of 0.5 or 1 (n=87).

**Table 1.** Demographic and clinical variables by diagnostic group.

|  | **Cognitively**<br>**Normal Older**<br>**Adults (CN)**<br>**(n= 276)** | **Mild Cognitive**<br>**Impairment**<br>**(MCI) (n=142)** | **Probable**<br>**Alzheimer's**<br>**Disease (AD) (n**<br>**= 87)** | **Group**<br>**Comparison**<br>***p*** |
| --- | --- | --- | --- | --- |
| **Age** | 70.84±7.95<br>55-87 | 73.54±6.81<br>55-90 | 73.13±8.53<br>55-89 | <0.001*<br>CN < MCI &<br>AD |
| **Sex (F)** | 51.5% | 66.2% | 66.7% | 0.003*<br>CN < MCI &<br>AD |
| **Years of**<br>**Education** | 12.10±4.74<br>0-25 | 10.13±4.41<br>0-20 | 9.85±5.39<br>0-20 | <0.001*<br>CN > MCI &<br>AD |
| ***APOE* ε4**<br>**allele**<br>**Positive** | 18.8% | 40.1% | 56.3% | <0.001*<br>CN < MCI <<br>AD |

### Image Acquisition and Processing

MR images were acquired on a Siemens 3T Biograph MMR scanner at Seoul National University, as previously described [39]. MR imaging included three-dimensional (3D)-T1-weighted magnetization-prepared rapid acquisition with gradient echo (MPRAGE) sequence scans obtained in sagittal orientation. The following parameters were used for 3D T1-weighted images: repetition time = 1670 ms, echo time = 1.89 ms, field of view = 250 mm, matrix size = 256 × 256, and slice thickness = 1.0 mm. Diffusion weighted images were acquired in the Anterior-Posterior phase encoding direction, 66 slices, repetition time 9500 ms, echo time 92.00 ms, slice thickness 2.0 mm, field of view = 230 mm, matrix 114×104, using 8 zero diffusion weighting and 60 directions at diffusion-weighting b=1000 s/mm^2^.

Diffusion-weighted and MPRAGE images for the KBASE cohort were preprocessed at the Indiana University School of Medicine using the Indiana University Connectivity Pipeline (https://github.com/IUSCA/IUSM-connectivity-pipeline) using a standard FSL-based workflow. Preprocessing steps included motion and eddy-current correction, with outlier detection and replacement with FSL EDDY [40, 41], followed by diffusion tensor model fitting with FSL DTIfit to generate scalar images (FA, MD, AxD, RD). Then using the FSL tract-based spatial statistics workflow [42], DTI scalar images were registered to the FMRIB_58_FA Montreal Neurological Institute (MNI) space template. Using the Johns Hopkins University ICBM-DTI-81 White Matter Labels Atlas [43], the median value for each scalar measure was extracted from targeted regions of interest (ROIs), specifically the left and right cingulum bundles of the hippocampus.

Amyloid PET images were collected as previously described [39], which was collected simultaneously as three-dimensional (3D) [^11^C]PiB-PET and 3D T1-weighted MRI using the 3.0T PET-MR scanner. After intravenous administration of ∼555 MBq of [^11^C]PiB (range, 450-610 MBq), a 30-min emission scan was obtained after a 40 min uptake period after injection. The PiB-PET data was collected in list mode and was reconstructed with routine corrections such as uniformity, UTE-based attenuation, and decay corrections into a 256×256 image matrix using iterative methods (6 iterations with 21 subsets).

PiB-PET images for the KBASE cohort were preprocessed with Statistical Parametric Mapping 12 (SPM12; https://www.fil.ion.ucl.ac.uk/spm/software/spm12/). First, 40-70min static PiB-PET images were created with motion correction between frames. Each participant’s static PiB-PET images were co-registered to each individual’s T1 structural image from the same visit. Next, voxel-based segmentation of the T1 images generated transformation matrices to normalize each T1 image to standard MNI space. The transformation matrices were then used to normalize the aligned static PiB-PET images to MNI space. Finally, normalized PiB-PET scans were intensity corrected into SUVR images using a cerebellar grey matter ROI from the Centiloid project [44] and smoothed with an 8mm full-width half maximum (FWHM) kernel. Mean SUVR was extracted from the MTL using an ROI generated from the combination of the entorhinal cortex, fusiform, parahippocampal gyrus, and temporal pole ROIs from mean Freesurfer version 6 segmentations/parcellations of 30 CN participants from the Alzheimer’s Disease Neuroimaging Initiative (ADNI-2).

### Neuropsychological Testing

Participants underwent comprehensive neuropsychological testing in accordance with a standardized protocol incorporating the CERAD-K neuropsychological battery, as previously described [39, 45]. The current analyses focused *a priori* on tests of cognition and mood: the Mini-Mental State Examination in the Korean Version of the CERAD Assessment (MMSE-KC), a comprehensive test of cognition which replaced reading and writing items with two judgement items due to the high rate of illiteracy in Korea [45, 46]; Trail Making Tests A and B (TMTA and TMTB); Digit Span Forward and Backward; Geriatric Depression Scale (GDS); Clinical Dementia Rating Sum of Boxes (CDR-SB); Subjective Memory Complaints Questionnaire (SMCQ) Total Memory Decline score; CERAD Word List Recall immediate and delay total scores; and Logical Memory immediate and delay total scores.

### Statistical Analysis

All statistical analyses were performed using SPSS Statistics (IBM Corp. Released 2021. IBM SPSS Statistics for Windows, Version 28.0. Armonk, NY: IBM Corp). Analysis of covariance (ANCOVA) with age at time of scan and sex as covariates was used to analyze between-group differences in DTI values by diagnosis, with DTI values as response and diagnosis as a factor predictor. Partial correlations controlling for age, sex, and education were used to examine the associations between DTI scalar values (FA, MD, AxD, RD) in the CBH and cognitive test scores. Partial correlations controlling for age, sex, and *APOE* ε4 allele positivity were used to determine the relationship between [^11^C]PiB-PET tracer uptake in the MTL, where the CBH is located, and DTI scalar values in the CBH. Pearson’s *r* correlation value was used to determine effect size and was interpreted using Cohen’s guidelines (small= 0.10, medium= 0.30, large = 0.50).

Significance thresholds in all Bonferroni-adjusted post-hoc T-tests employed a *p*-value of 0.05. To control for multiple comparisons in partial correlations, significance was determined at a Bonferroni-corrected alpha-level. Significance of association between CBH DTI metrics and cognition (*n=*32 [12 tests per hemisphere and 4 scalar measures per hemisphere) was determined at the Bonferroni corrected *p*-value ≤ 0.0015. Significance for partial correlations between PET values and DTI (*n*= 8 [four tests per hemisphere]) was determined at the Bonferroni corrected *p*-value ≤ 0.00625.

## RESULTS

### CBH Differences Across the AD Continuum in KBASE

ANCOVA analysis results can be found in **Table 2**. FA values were significantly lower and RD, MD, and AxD values were significantly higher (all *p* < 0.001) in MCI and AD subjects compared to CN, indicating a significant difference of WM integrity in the bilateral CBH between diagnostic group after controlling for age and sex.

**Table 2.** Overall and post-hoc between-group differences of WM integrity in the left and right cingulum bundle of the hippocampus (CBH) as measured through the 4 commonly acquired diffusion tensor imaging (DTI) indices: fractional anisotropy (FA), radial diffusivity (RD), axial diffusivity (AxD), and mean diffusivity (MD). When controlling for age and sex, all groups had significantly different average DTI values in each hemisphere. CN = cognitively normal; MCI = mild cognitive impairment; AD = probable Alzheimer’s disease-related dementia. Overall *p* significance determined at the Bonferroni-adjusted alpha level of 0.0015, and automatically Bonferroni-adjusted post-hoc T-tests employed a *p*-value of 0.05.

| ANCOVA |  |  |  |  |  |  |  |  |
| --- | --- | --- | --- | --- | --- | --- | --- | --- |
|  | Left Cingulum Bundle of the Hippocampus |  |  |  | Right Cingulum Bundle of the Hippocampus |  |  |  |
| | $F(2,500)$ | $p$ | $\eta_p^2$ | Post-hoc | $F(2,500)$ | $p$ | $\eta_p^2$ | Post-hoc |
| **FA** | 61.562 | <0.001 | 0.198 | CN and MCI: $p = 0.002$<br>CN and AD: $p < 0.001$<br>MCI and AD: $p < 0.001$ | 31.359 | <0.001 | 0.111 | CN and MCI: $p = 0.015$<br>CN and AD: $p < 0.001$<br>MCI and AD: $p < 0.001$ |
| **RD** | 80.705 | <0.001 | 0.244 | CN and MCI: $p < 0.001$<br>CN and AD: $p < 0.001$<br>MCI and AD: $p < 0.001$ | 96.300 | <0.001 | 0.278 | CN and MCI: $p < 0.001$<br>CN and AD: $p < 0.001$<br>MCI and AD: $p < 0.001$ |
| **AxD** | 39.835 | <0.001 | 0.137 | CN and MCI: $p < 0.001$<br>CN and AD: $p < 0.001$<br>MCI and AD: $p < 0.001$ | 82.884 | <0.001 | 0.249 | CN and MCI: $p < 0.001$<br>CN and AD: $p < 0.001$<br>MCI and AD: $p < 0.001$ |
| **MD** | 81.016 | <0.001 | 0.245 | CN and MCI: $p < 0.001$<br>CN and AD: $p < 0.001$<br>MCI and AD: $p < 0.001$ | 110.980 | <0.001 | 0.307 | CN and MCI: $p < 0.001$<br>CN and AD: $p < 0.001$<br>MCI and AD: $p < 0.001$ |

Diagnostic group differences are shown in ***Figure 1 a-h***. Bonferroni-adjusted post-hoc tests revealed that CN participants had significantly lower RD, MD, and AxD values than MCI (all p-values <0.001). CN participants also had significantly higher FA than MCI (left *p* = 0.002, right *p*=0.015). Though average MCI DTI values reflected lower WM integrity than CN, AD values reflected lower WM integrity than both MCI and CN (all *p*-values < 0.001).

**Figure 1.**
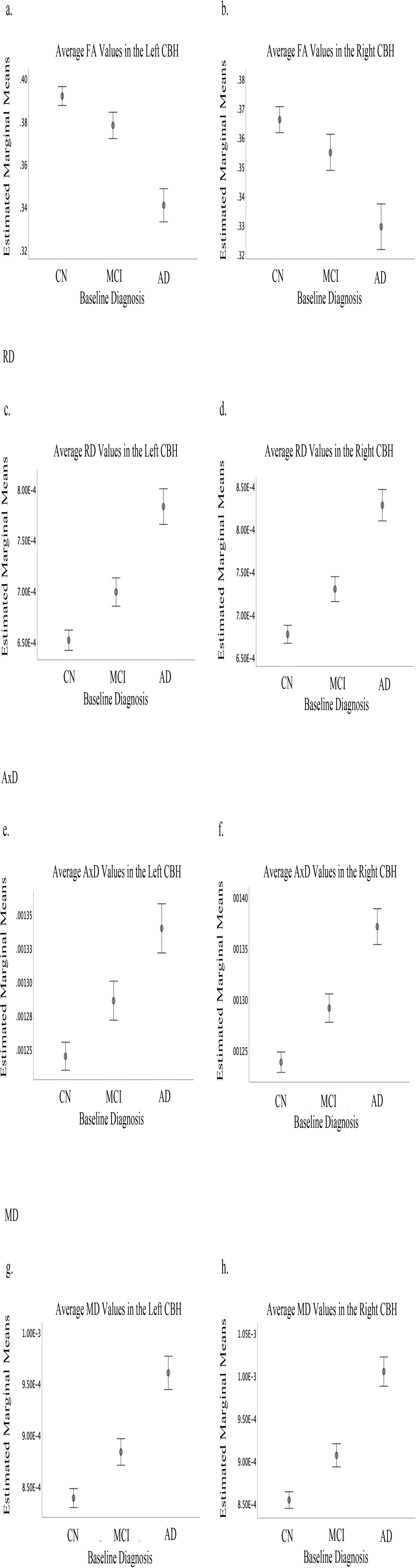
Differences in WM integrity metrics by diagnostic group. Estimated marginal mean per group (CN = cognitively normal; MCI = mild cognitive impairment; AD = probable Alzheimer’s disease dementia) of left and right cingulum bundles of the hippocampus (CBH) values per each diffusion tensor imaging (DTI) scalar measure (FA = Fractional Anisotropy; RD = Radial Diffusivity; AxD = Axial Diffusivity; MD = Mean Diffusivity), controlled for age and sex. Error bars represent 95% confidence interval. All groups demonstrated significantly different average values for DTI scalar measures, with MCI values falling intermediately between CN and AD values.

### Relationship of CBH DTI values to Cognition

Associations between cognitive tests and CBH WM integrity indices can be found in **Table 3**. Decreased WM integrity in the CBH, as reflected by lower FA and higher RD, AxD, and MD, was significantly associated with poorer cognition, as well as more depressed mood. Specifically, significant associations were observed before adjustment for multiple comparisons between WM integrity and performance on the Digit Span Backward, TMTA, and TMTB, MMSE-KC, CDR-SB, Word List Recall scores, Logical Memory scores, and GDS (all *p*-values ≤ 0.003) with small to moderate effect sizes (|*r*| *=* 0.131 – 0.543). Of these, the smallest effect size was seen between AxD values and Digit Span Backward in the left CBH (*r* = −0.131, *p* = 0.003), which did not meet significance at the Bonferroni-adjusted alpha-level of 0.0015, and the largest effect size was seen between GDS scores and MD values in the right CBH (*r* = 0.543, *p* < 0.001). Better Digit Span Forward scores were not associated with FA or RD values, though prior to adjustment for multiple comparisons, performance was negatively associated with lower bilateral AxD values as well as MD values in the left CBH only (*p*≤0.004) with small effect size. At the Bonferroni-adjusted alpha-level, only AxD values of the left CBH were significantly associated with Digit Span Forward scores. Finally, more memory complaints (higher total SMCQ score) were significantly associated with higher bilateral RD, AxD and MD values (*r* = 0.152 - 0.201, all *p* < 0.001), though not with FA.

**Table 3.** Association of DTI values from the left (a) and right (b) CBH with cognitive exams. Significance is determined at the Bonferroni-adjusted alpha level of 0.0015 and all partial correlations in this table are controlled for age, sex, and years of education at time of scan. TMTA = Trail Making Test A; TMTB = Trail Making Test B; MMSE-KC = Mini-Mental State Exam in the Korean Version of the CERAD; CDR = Clinical Dementia Rating Scale; SMCQ = Subjective Memory Complaints Questionnaire.

| **a. Association of Imaging Measures with Cognitive Exams in the Left Cingulum Bundle of the Hippocampus** |  |  |  |  |  |  |  |  |
| --- | --- | --- | --- | --- | --- | --- | --- | --- |
| **Cognitive Measure** | **FA** |  | **RD** |  | **AxD** |  | **MD** |  |
|  | *r* | *p* | *r* | *p* | *r* | *p* | *r* | *p* |
| Digit Span Forward | 0.052 | 0.249 | -0.115 | 0.010 | -0.194 | <0.001* | -0.129 | 0.004 |
| Digit Span Backward | 0.196 | <0.001* | -0.172 | <0.001* | -0.131 | 0.003 | -0.146 | 0.001* |
| TMTA | -0.246 | <0.001* | 0.264 | <0.001* | 0.188 | <0.001* | 0.254 | <0.001* |
| TMTB | -0.195 | <0.001* | 0.226 | <0.001* | 0.175 | <0.001* | 0.217 | <0.001* |
| Geriatric Depression Scale | -0.445 | <0.001* | 0.486 | <0.001* | 0.363 | <0.001* | 0.485 | <0.001* |
| MMSE-KC | 0.395 | <0.001* | -0.462 | <0.001* | -0.350 | <0.001* | -0.464 | <0.001* |
| CDR Sum of Boxes | -0.445 | <0.001* | 0.484 | <0.001* | 0.345 | <0.001* | 0.478 | <0.001* |
| SMCQ Total Memory Decline | -0.097 | 0.030 | 0.166 | <0.001* | 0.152 | <0.001* | 0.176 | <0.001* |
| Word List Recall (Immediate) Total Score | 0.230 | <0.001* | -0.282 | <0.001* | -0.270 | <0.001* | -0.293 | <0.001* |
| Word List Recall (Delay) Total Score | 0.405 | <0.001* | -0.457 | <0.001* | -0.353 | <0.001* | -0.461 | <0.001* |
| Logical Memory (Immediate) Total Score | 0.356 | <0.001* | -0.385 | <0.001* | -0.287 | <0.001* | -0.382 | <0.001* |
| Logical Memory (Delay) Total Score | 0.321 | <0.001* | -0.336 | <0.001* | -0.240 | <0.001* | -0.327 | <0.001* |
| **b. Association of Imaging Measures with Cognitive Exams in the Right Cingulum Bundle of the Hippocampus** |  |  |  |  |  |  |  |  |
| **Cognitive Measure** | **FA** |  | **RD** |  | **AxD** |  | **MD** |  |
|  | *r* | *p* | *r* | *p* | *r* | *p* | *r* | *p* |
| Digit Span Forward | 0.041 | 0.358 | -0.098 | 0.028 | -0.136 | 0.002 | -0.098 | 0.029 |
| Digit Span Backward | 0.169 | <0.001* | -0.196 | <0.001* | -0.174 | <0.001* | -0.185 | <0.001* |
| TMTA | -0.291 | <0.001* | 0.382 | <0.001* | 0.305 | <0.001* | 0.386 | <0.001* |
| TMTB | -0.188 | <0.001* | 0.252 | <0.001* | 0.271 | <0.001* | 0.261 | <0.001* |
| Geriatric Depression Scale | -0.327 | <0.001* | 0.520 | <0.001* | 0.488 | <0.001* | 0.543 | <0.001* |
| MMSE-KC | 0.332 | <0.001* | -0.501 | <0.001* | -0.461 | <0.001* | -0.517 | <0.001* |
| CDR Sum of Boxes | -0.334 | <0.001* | 0.516 | <0.001* | 0.470 | <0.001* | 0.538 | <0.001* |
| SMCQ Total Memory Decline | -0.070 | 0.117 | 0.187 | <0.001* | 0.200 | <0.001* | 0.201 | <0.001* |
| Word List Recall (Immediate) Total Score | 0.206 | <0.001* | -0.288 | <0.001* | -0.288 | <0.001* | -0.292 | <0.001* |
| Word List Recall (Delay) Total Score | 0.313 | <0.001* | -0.473 | <0.001* | -0.441 | <0.001* | -0.485 | <0.001* |
| Logical Memory (Immediate) Total Score | 0.285 | <0.001* | -0.383 | <0.001* | -0.361 | <0.001* | -0.392 | <0.001* |
| Logical Memory (Delay) Total Score | 0.278 | <0.001* | -0.363 | <0.001* | -0.325 | <0.001* | -0.364 | <0.001* |

### Relationship of CBH WM Integrity Values to [^*11*^*C]PiB-PET Values*

Associations between average [^11^C]PiB-PET SUVR values in the MTL and CBH WM integrity measures can be found in **Table 4**. As observed in **Figure 2**, average SUVR values in the MTL were significantly different between all three diagnostic groups in a stepwise fashion (*p* <0.001, η_p_^2^= 0.222) with significantly lower SUVR values in CN participants than MCI, and lower in MCI than AD, reflecting increasing amyloid burden across the disease continuum (all *p*-values < 0.001). Poorer WM integrity in the CBH, as reflected by lower FA and higher RD, AxD, and MD, was significantly associated with more amyloid burden in the MTL as reflected by [^11^C]PiB-PET SUVR. Bilaterally, higher FA values were negatively correlated and lower RD, AxD and MD values were positively correlated with SUVR in the MTL (|*r*| = 0.233 – 0.330, *p*< 0.001).

**Table 4.** Association of WM integrity metrics with amyloid deposition. Pearson’s partial correlations between average [^11^C]PiB-PET SUVR in the medial temporal lobe (MTL) and DTI indices in the left and right CBH. Significance is determined at the Bonferroni-adjusted p-value of 0.00625 and all partial correlations in this table are controlled for age, sex, and APOE ε4 allele positivity. All associations were significant (p <0.001) with small to medium effect sizes.

| Partial Correlations between [$^{11}\text{C}$]PiB PET Values of the Medial Temporal Lobe and DTI in the CBH | | | | |
| --- | --- | --- | --- | --- |
|  | Left Cingulum Bundle of the Hippocampus |  | Right Cingulum Bundle of the Hippocampus |  |
|  | *r* | *p* | *r* | *p* |
| ***FA*** | -0.267 | <0.001* | -0.233 | <0.001* |
| ***RD*** | 0.330 | <0.001* | 0.325 | <0.001* |
| ***AxD*** | 0.240 | <0.001* | 0.279 | <0.001* |
| ***MD*** | 0.323 | <0.001* | 0.323 | <0.001* |

**Figure 2:**
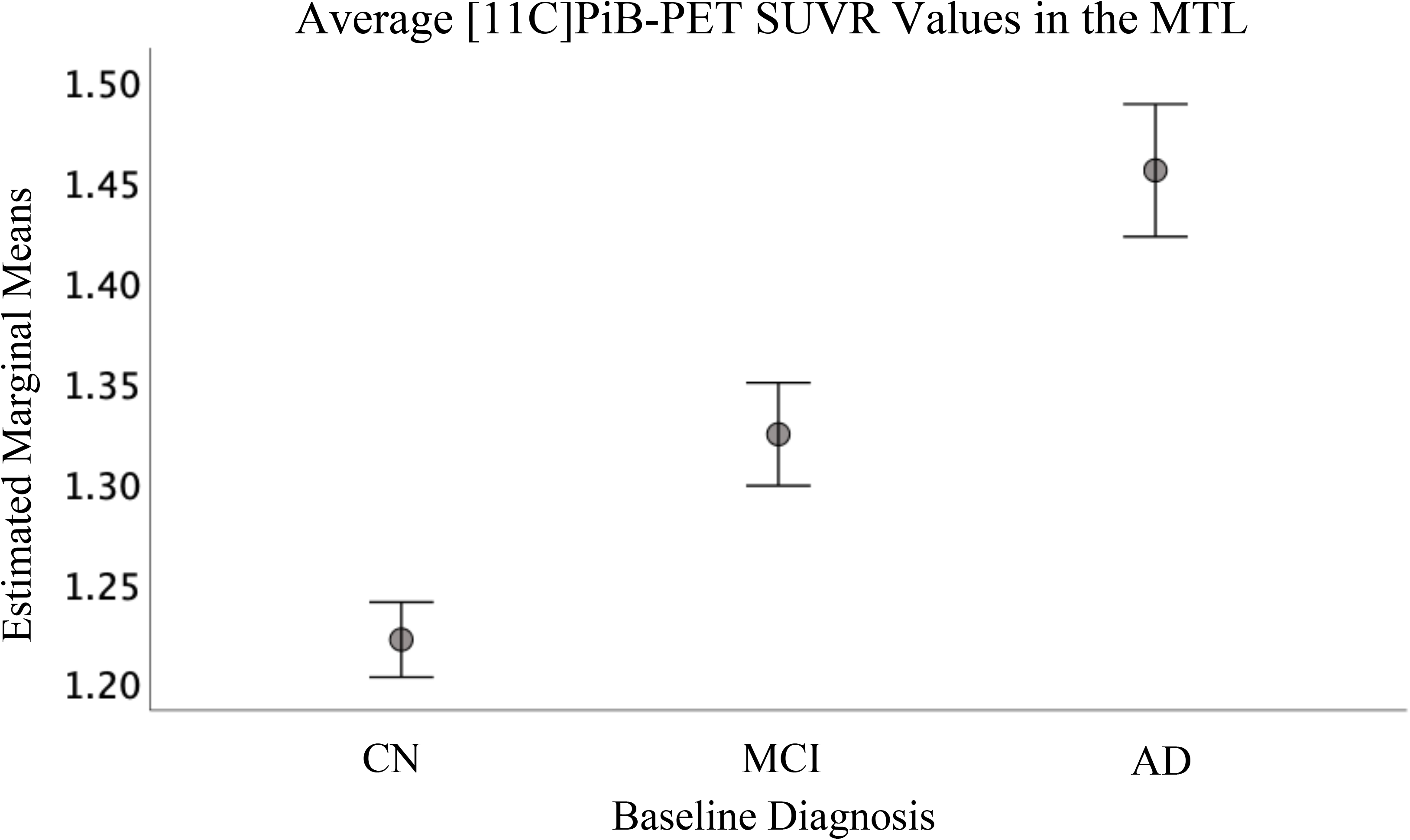
Medial temporal amyloid deposition by diagnostic group. Estimated marginal mean per group (CN, MCI, AD) of the medial temporal lobe, which includes the Papez circuit, cingulum bundle of the hippocampus (CBH) as well as other circuits relevant to AD. All three groups demonstrate significantly different average SUVR values after controlling for age, sex, and APOE e4 allele positivity (F(2,499) = 71.104, p <0.001, η_p_^2^= 0.222), with error bars representing 95% confidence interval. As seen in DTI values, participants with MCI demonstrate an intermediate SUVR uptake that is significantly higher than CN, but less than AD participants.

## DISCUSSION

The CBH is part of the cingulum, which is a tract that is critically involved in interconnection of the frontal, medial temporal, and parietal cortices [11, 12], as well as in the function of the limbic Papez Circuit[12]. Previously, disrupted integrity of this circuit, especially within the parahippocampal cingulum, has been demonstrated in AD [13, 14, 25, 27-34]. However, these investigations have been limited to predominantly NHW populations, which affects the broad generalizability of this structure’s role in AD. Here, we report reduced WM integrity of the CBH in Korean AD and MCI subjects compared to CN, as well as associations between lower WM integrity and poorer cognition and lower WM integrity and higher amyloid burden in a cohort from the initial phase of KBASE. These results provide evidence of a role for the CBH in participants on the AD continuum with Korean ancestry and are consistent with prior literature, which has established this relationship in cohorts of predominantly NHW ancestry [27-34].

Previously, higher amyloid burden as observed through PET has been associated with lower FA in CN subjects at risk for AD [35]. Our study extended these results, demonstrating further association between amyloid burden and the other scalar DTI measures (RD, AxD, and MD). Despite worsened WM integrity being associated with higher amyloid burden and thus implied disease progression, we are unable conclude a specific temporal association between amyloid-beta plaque deposition and poorer WM integrity, nor can we determine whether these are independent or intertwined processes. However, the significant differences in diffusion indices in the CBH between CN, MCI, and AD participants, with MCI values falling intermediately between CN and AD **(Figure 1)**, mirroring PiB tracer uptake in the MTL **(Figure 2)**, indicate the possibility that this region may be a sensitive measure for diagnostic staging.

Moreover, most neuropsychological test scores were significantly associated with all four CBH WM integrity values. The Digit Span Forward, which tests the domain of working memory that is often impaired in AD, was less robustly associated with poorer WM integrity. Presently, there is not a consensus in the literature regarding the relationship between the cingulum and working memory. Working memory seems to be relatively spared in the event of damage to the cingulum [11]. For example, recent evidence has shown that some tests of working memory were not associated with diffusion values in the cingulum in multiple sclerosis, a neurodegenerative disease involving myelin [47], although other studies have suggested a relationship between lower cingulum bundle integrity and poorer working memory in cognitively normal adults [48, 49]. The association prior to multiple comparison correction which was seen in the present study between Digit Span Forward and AxD values could potentially indicate a specific role for axonal degeneration, as it is thought that AxD is more specific to axonal injury than other scalar measures [50]. The Digit Span Backward is a similar test which also involves working memory, though incorporates executive functioning skills as well, which may explain the higher associations seen here between worsening WM integrity and poorer scores compared to Digit Span Forward. Additionally, SMCQ Total Memory Decline, a subjective test, was not strongly associated with FA. Patients with mild memory complaints who are filling out the exam for themselves may not be fully cognizant of the extent of their disease, or stigma may prevent older adults from fully expressing their concerns [4, 5, 38], though further studies are needed to understand why FA values, which typically are myelin-driven but can reflect axonal damage[26], are less sensitive.

Additionally, while AxD is specific to axonal degeneration, RD is thought to correspond more specifically to myelin-specific damage [51]. MD, like FA, may also partially indicate myelin-related pathology [52]. Here, we found that the GDS was strongly associated with WM integrity in the bilateral CBH, with the strongest effect sizes seen in RD and MD. It is possible that depression, previously identified in NHW cohorts [21, 22], as well as Korean [23], as a major risk factor for AD, may be related to myelin degeneration within the limbic system and specifically the CBH.

Limitations of this study include the considerably smaller AD subgroup compared to MCI and CN groups, which may reduce statistical power. Additionally, the diagnosis of AD is probable and cannot be proven until post-mortem examination, though all probable AD-dementia participants were thoroughly cognitively tested and met established guidelines. Finally, while DTI is considered an advanced neuroimaging metric, it still requires further validation to confirm that it is definitively and accurately representative of WM integrity. Future work is needed to fully characterize the pathological differences of the CBH in relation to tau and amyloid, as well as to fully characterize this region in large cohorts of other underrepresented minorities on the AD continuum, such as African American, Hispanic, and other minority communities.

In conclusion, we demonstrate that loss of WM integrity in the CBH in a cohort of older adult Koreans on the AD continuum is consistent with the patterns seen in predominately NHW cohorts. Worsening WM integrity in this region was associated with MCI and AD diagnosis, as well as poorer cognition in Koreans. Poorer WM integrity in this region is also associated with higher scores on the Geriatric Depression Scale, supporting depression as an AD risk factor in both White and Korean populations. Finally, worsening WM integrity is associated with higher amyloid burden, demonstrating a similar pattern of progression across the AD continuum. In sum, the CBH is a promising region that may be sensitive to MCI and AD diagnoses across multiple racial/ethnic populations and should be further investigated in diverse cohorts.

## Data Availability

All data produced in the present study are available upon reasonable request to the authors.

## ACKNOWLEDGEMENTS

The authors would like to thank Dr. Michelle Block and Matt Tharp for valuable discussions, as well as Dr. Paula J. Bice for editorial assistance.

## CONSENT STATEMENT

The cohort was chosen from data collected as part of the initial phase of the Korean Brain Aging Study for the Early Diagnosis and Prediction of Alzheimer’s Disease (KBASE) with approval by the Institutional Review Boards of Seoul National University (SNU) Hospital and SNU-SMG Boramae Medical Center and all human subjects provided informed consent in accordance with the Declaration of Helsinki as previously described [39].

## REFERENCES

[1] C. R. Jack et al., “A/T/N: an unbiased descriptive classification scheme for Alzheimer disease biomarkers,” Neurology, vol. 87, no. 5, pp. 539–547, 2016.

[2] H. Braak and E. Braak, “Neuropathological stageing of Alzheimer-related changes,” Acta neuropathologica, vol. 82, no. 4, pp. 239–259, 1991.

[3] P. S. Aisen et al., “On the path to 2025: understanding the Alzheimer’s disease continuum,” Alzheimer’s research & therapy, vol. 9, no. 1, pp. 1–10, 2017.

[4] K. N. Fargo, M. C. Carrillo, M. W. Weiner, W. Z. Potter, and Z. Khachaturian, “The crisis in recruitment for clinical trials in Alzheimer’s and dementia: An action plan for solutions,” Alzheimer’s & dementia: the journal of the Alzheimer’s Association, vol. 12, no. 11, pp. 1113–1115, 2016.

[5] A. L. Gilmore-Bykovskyi et al., “Recruitment and retention of underrepresented populations in Alzheimer’s disease research: a systematic review,” Alzheimer’s & Dementia: Translational Research & Clinical Interventions, vol. 5, pp. 751–770, 2019.

[6] E. Kornblith, A. Bahorik, W. J. Boscardin, F. Xia, D. E. Barnes, and K. Yaffe, “Association of race and ethnicity with incidence of dementia among older adults,” JAMA, vol. 327, no. 15, pp. 1488–1495, 2022.

[7] A. Miyashita, M. Kikuchi, N. Hara, and T. Ikeuchi, “Genetics of Alzheimer’s disease: an East Asian perspective,” Journal of Human Genetics, pp. 1–10, 2022.

[8] L. R. Hirschfeld, S. L. Risacher, K. Nho, and A. J. Saykin, “Myelin repair in Alzheimer’s disease: a review of biological pathways and potential therapeutics,” Translational Neurodegeneration, vol. 11, no. 1, p. 47, d2022/10/26 2022, doi: 10.1186/s40035-022-00321-1.

[9] G. Bartzokis, “Alzheimer’s disease as homeostatic responses to age-related myelin breakdown,” Neurobiology of aging, vol. 32, no. 8, pp. 1341–1371, 2011.

[10] G. Bartzokis, P. H. Lu, and J. Mintz, “Human brain myelination and amyloid beta deposition in Alzheimer’s disease,” Alzheimer’s & Dementia, vol. 3, no. 2, pp. 122-125, 2007/04/01/ 2007, doi: https://doi.org/10.1016/j.jalz.2007.01.019.

[11] E. J. Bubb, C. Metzler-Baddeley, and J. P. Aggleton, “The cingulum bundle: anatomy, function, and dysfunction,” Neuroscience & Biobehavioral Reviews, vol. 92, pp. 104–127, 2018.

[12] G. Forno, A. Lladó, and M. Hornberger, “Going round in circles—The Papez circuit in Alzheimer’s disease,” European Journal of Neuroscience, vol. 54, no. 10, pp. 7668–7687, 2021.

[13] W. Li et al., “Aberrant functional connectivity in Papez circuit correlates with memory performance in cognitively intact middle-aged APOE4 carriers,” Cortex, vol. 57, pp. 167–176, 2014.

[14] A. Pichet Binette et al., “Bundle-specific associations between white matter microstructure and Aβ and tau pathology in preclinical Alzheimer’s disease,” Elife, vol. 10, p. e62929, 2021.

[15] M. Van Den Heuvel, R. Mandl, J. Luigjes, and H. H. Pol, “Microstructural organization of the cingulum tract and the level of default mode functional connectivity,” Journal of Neuroscience, vol. 28, no. 43, pp. 10844–10851, 2008.

[16] K. Mevel, G. Chételat, F. Eustache, and B. Desgranges, “The default mode network in healthy aging and Alzheimer’s disease,” International journal of Alzheimer’s disease, vol. 2011, 2011.

[17] M. Yu, O. Sporns, and A. J. Saykin, “The human connectome in Alzheimer disease— relationship to biomarkers and genetics,” Nature Reviews Neurology, vol. 17, no. 9, pp. 545–563, 2021.

[18] K. D. Bhatia, L. A. Henderson, E. Hsu, and M. Yim, “Reduced integrity of the uncinate fasciculus and cingulum in depression: a stem-by-stem analysis,” Journal of Affective Disorders, vol. 235, pp. 220–228, 2018.

[19] N. Mertse et al., “Associations between anterior cingulate thickness, cingulum bundle microstructure, melancholia and depression severity in unipolar depression,” Journal of affective disorders, vol. 301, pp. 437–444, 2022.

[20] M. Li, F. Wu, Y. Cao, X. Jiang, L. Kong, and Y. Tang, “Abnormal white matter integrity in Papez circuit in first-episode medication-naive adults with anxious depression: A combined voxel-based analysis and region of interest study,” Journal of Affective Disorders, 2023.

[21] S. Van der Mussele et al., “Prevalence and associated behavioral symptoms of depression in mild cognitive impairment and dementia due to Alzheimer’s disease,” International Journal of Geriatric Psychiatry, vol. 28, no. 9, pp. 947–958, 2013.

[22] B. S. Diniz, M. A. Butters, S. M. Albert, M. A. Dew, and C. F. Reynolds, “Late-life depression and risk of vascular dementia and Alzheimer’s disease: systematic review and meta-analysis of community-based cohort studies,” The British Journal of Psychiatry, vol. 202, no. 5, pp. 329–335, 2013.

[23] J. B. Bae et al., “Incidence of and risk factors for Alzheimer’s disease and mild cognitive impairment in Korean elderly,” Dementia and geriatric cognitive disorders, vol. 39, no. 1-2, pp. 105–115, 2015.

[24] Y. Wu, D. Sun, Y. Wang, Y. Wang, and S. Ou, “Segmentation of the cingulum bundle in the human brain: a new perspective based on DSI tractography and fiber dissection study,” Frontiers in neuroanatomy, vol. 10, p. 84, 2016.

[25] I. A. Clark, S. Mohammadi, M. F. Callaghan, and E. A. Maguire, “Conduction velocity along a key white matter tract is associated with autobiographical memory recall ability,” Elife, vol. 11, p. e79303, 2022.

[26] J. Soares, P. Marques, V. Alves, and N. Sousa, “A hitchhiker’s guide to diffusion tensor imaging,” Frontiers in neuroscience, vol. 7, p. 31, 2013.

[27] Y. Wang et al., “Selective changes in white matter integrity in MCI and older adults with cognitive complaints,” Biochimica et Biophysica Acta (BBA)-Molecular Basis of Disease, vol. 1822, no. 3, pp. 423–430, 2012.

[28] Q. Wen et al., “White matter alterations in early-stage Alzheimer’s disease: A tract-specific study,” Alzheimer’s & Dementia: Diagnosis, Assessment & Disease Monitoring, vol. 11, pp. 576-587, 2019/12/01/ 2019, doi: https://doi.org/10.1016/j.dadm.2019.06.003.

[29] I. H. Choo et al., “Posterior cingulate cortex atrophy and regional cingulum disruption in mild cognitive impairment and Alzheimer’s disease,” Neurobiology of aging, vol. 31, no. 5, pp. 772–779, 2010.

[30] E. Gozdas, H. Fingerhut, L. C. Chromik, R. O’Hara, A. L. Reiss, and S. Hosseini, “Focal white matter disruptions along the cingulum tract explain cognitive decline in amnestic mild cognitive impairment (aMCI),” Scientific reports, vol. 10, no. 1, pp. 1–10, 2020.

[31] Y. Liu et al., “Diffusion tensor imaging and tract-based spatial statistics in Alzheimer’s disease and mild cognitive impairment,” Neurobiology of aging, vol. 32, no. 9, pp. 1558–1571, 2011.

[32] Y. Zhang et al., “Diffusion tensor imaging of cingulum fibers in mild cognitive impairment and Alzheimer disease,” Neurology, vol. 68, no. 1, p. 13–19, 2007.

[33] J. L. Dalboni da Rocha, I. Bramati, G. Coutinho, F. Tovar Moll, and R. Sitaram, “Fractional Anisotropy changes in parahippocampal cingulum due to Alzheimer’s Disease,” Scientific reports, vol. 10, no. 1, p. 1–8, 2020.

[34] A. Zavaliangos-Petropulu et al., “Diffusion MRI indices and their relation to cognitive impairment in brain aging: the updated multi-protocol approach in ADNI3,” Frontiers in Neuroinformatics, vol. 13, p. 2, 2019.

[35] A. Rieckmann, K. R. Van Dijk, R. A. Sperling, K. A. Johnson, R. L. Buckner, and T. Hedden, “Accelerated decline in white matter integrity in clinically normal individuals at risk for Alzheimer’s disease,” Neurobiology of aging, vol. 42, p. 177–188, 2016.

[36] Y.-C. Lin et al., “Cingulum correlates of cognitive functions in patients with mild cognitive impairment and early Alzheimer’s disease: a diffusion spectrum imaging study,” Brain topography, vol. 27, p. 393–402, 2014.

[37] P. R. Mena et al., “The Alzheimer’s Disease Sequencing Project–Follow Up Study (ADSPLJFUS): Increasing ethnic diversity in Alzheimer’s genetics research with the addition of potential new cohorts,” Alzheimer’s & Dementia, vol. 17, p. e056101, 2021.

[38] A. L. Chin, S. Negash, and R. Hamilton, “Diversity and disparity in dementia: The impact of ethnoracial differences in Alzheimer’s disease,” Alzheimer disease and associated disorders, vol. 25, no. 3, p. 187, 2011.

[39] M. S. Byun et al., “Korean brain aging study for the early diagnosis and prediction of Alzheimer’s disease: methodology and baseline sample characteristics,” Psychiatry investigation, vol. 14, no. 6, p. 851, 2017.

[40] J. L. Andersson and S. N. Sotiropoulos, “An integrated approach to correction for off-resonance effects and subject movement in diffusion MR imaging,” Neuroimage, vol. 125, p. 1063–1078, 2016.

[41] J. L. Andersson, M. S. Graham, E. Zsoldos, and S. N. Sotiropoulos, “Incorporating outlier detection and replacement into a non-parametric framework for movement and distortion correction of diffusion MR images,” Neuroimage, vol. 141, p. 556–572, 2016.

[42] S. M. Smith et al., “Tract-based spatial statistics: voxelwise analysis of multi-subject diffusion data,” Neuroimage, vol. 31, no. 4, p. 1487–1505, 2006.

[43] S. Mori, S. Wakana, P. C. Van Zijl, and L. Nagae-Poetscher, MRI atlas of human white matter. Elsevier, 2005.

[44] W. E. Klunk et al., “The Centiloid Project: standardizing quantitative amyloid plaque estimation by PET,” Alzheimer’s & dementia : the journal of the Alzheimer’s Association, vol. 11, no. 1, p. 1-15 e1-4, Jan 2015, doi: 10.1016/j.jalz.2014.07.003.

[45] D. Y. Lee et al., “A normative study of the CERAD neuropsychological assessment battery in the Korean elderly,” Journal of the International Neuropsychological Society, vol. 10, no. 1, p. 72–81, 2004.

[46] D.-Y. Lee et al., “A normative study of the mini-mental state examination in the Korean elderly,” Journal of Korean Neuropsychiatric Association, p. 508–525, 2002.

[47] K. A. Koenig et al., “The relationship between cognitive function and high-resolution diffusion tensor MRI of the cingulum bundle in multiple sclerosis,” Multiple Sclerosis Journal, vol. 21, no. 14, p. 1794–1801, 2015.

[48] M. Takahashi, K. Iwamoto, H. Fukatsu, S. Naganawa, T. Iidaka, and N. Ozaki, “White matter microstructure of the cingulum and cerebellar peduncle is related to sustained attention and working memory: a diffusion tensor imaging study,” Neuroscience letters, vol. 477, no. 2, p. 72–76, 2010.

[49] R. A. Charlton, T. R. Barrick, I. N. C. Lawes, H. S. Markus, and R. G. Morris, “White matter pathways associated with working memory in normal aging,” Cortex, vol. 46, no. 4, p. 474–489, 2010.

[50] M. D. Budde, M. Xie, A. H. Cross, and S.-K. Song, “Axial diffusivity is the primary correlate of axonal injury in the experimental autoimmune encephalomyelitis spinal cord: a quantitative pixelwise analysis,” Journal of Neuroscience, vol. 29, no. 9, p. 2805–2813, 2009.

[51] S.-K. Song, S.-W. Sun, M. J. Ramsbottom, C. Chang, J. Russell, and A. H. Cross, “Dysmyelination revealed through MRI as increased radial (but unchanged axial) diffusion of water,” Neuroimage, vol. 17, no. 3, p. 1429–1436, 2002.

[52] J. M. Peters et al., “White matter mean diffusivity correlates with myelination in tuberous sclerosis complex,” Annals of clinical and translational neurology, vol. 6, no. 7, p. 1178–1190, 2019.

